# Development of a Rapid Automated Point-of-Care Test for *Mycobacterium tuberculosis* Detection from Tongue Swabs and Sputum Specimens on the DASH® Rapid PCR System

**DOI:** 10.64898/2026.02.26.26347105

**Authors:** Matthew A. Butzler, Jennifer L. Reed, Alaina M. Olson, Rachel C. Wood, Gerard A. Cangelosi, Angelique K. Luabeya, Mark Hatherill, Arthur M. Chiwaya, Loren Rockman, Grant Theron, Sally M. McFall

**Affiliations:** Center for Innovation in Global Health Technologies, Robert J. Havey Institute for Global Health, Northwestern University Feinberg School of Medicine, Chicago, Illinois, 60611, USA; Department of Biomedical Engineering, Northwestern University, Evanston, IL 60208, USA; Department of Environmental and Occupational Health Sciences, University of Washington, Seattle, Washington, USA; South African Tuberculosis Vaccine Initiative, Institute of Infectious Disease & Molecular Medicine and Department of Pathology, University of Cape Town, Cape Town, South Africa; DSI-NRF Centre of Excellence for Biomedical Tuberculosis Research, South African Medical Research Council Centre for Tuberculosis Research, Division of Molecular Biology and Human Genetics, Faculty of Medicine and Health Sciences, Stellenbosch University, South Africa

## Abstract

*Mycobacterium tuberculosis* (MTB) disease is a major global health threat with most tuberculosis (TB) cases occurring in low-and middle-income countries (LMIC) with limited healthcare infrastructure. Near-point-of-care testing which can be deployed at peripheral clinical settings is needed to start treatment earlier and thereby improve treatment outcomes. Here we report the development and preliminary characterization of an MTB detection assay that utilizes tongue swab or sputum specimens for The DASH® Rapid PCR System which employs cartridge-based automated sequence specific capture sample prep combined with dual target qPCR multicopy MTB insertion sequences IS*6110* and IS*1081* amplification and detection. MTB is resistant to conventional bacterial lysis techniques; therefore, we evaluated two pre-cartridge lysing techniques, mechanical lysis and sonication, and selected sonication for all subsequent studies. The DASH MTB assay demonstrated a limit of detection of 2.5 MTB cells/swab with no detection of 10 non-tuberculosis *Mycobacterium* strains. Clinical testing of 100 (49 positive and 51 negative) de-identified blinded sputa from South African symptomatic clinic attendees yielded an overall test sensitivity of 96% (100% for smear positive samples and 88% for smear negative samples) and specificity of 88% when compared to sputum culture. In a separate study of 110 tongue swab specimens (70 positive and 40 negative) from South African symptomatic clinic attendees, the sensitivity was 91% and the specificity was 100%. We further demonstrated that the test is compatible with peripheral LMIC settings via external battery operation and cartridge stability at 45°C for up to one year.

**Importance:** Tuberculosis (TB) is the single most deadly infectious disease with 1.23 million deaths in 2024. Near-point-of-care testing which can be deployed at peripheral settings that lack laboratory infrastructure to deliver prompt and accurate diagnosis is needed to start treatment earlier and thereby improve treatment outcomes. In this study, we have developed an automated test to detect *Mycobacterium tuberculosis* (MTB), the cause of TB, from sputum and tongue swab specimens. Its high sensitivity and specificity, rapid time to result, and compatibility with environments that lack air conditioning and consistent electricity make this assay suitable for diverse clinical settings.

## Introduction

Tuberculosis (TB) is the leading cause of death from a single infectious disease with more than 10 million people contracting TB resulting in 1.23 million deaths in 2024 with most deaths occurring in low-and-middle income countries (1, 2). Delayed and missed diagnosis is a major impediment in providing TB care (3) . Centralized testing can lead to extensive delays in results and loss to follow up in the care cascade. Near-point-of-care (nPOC) testing using instruments that are preferably battery operated and do not require specialized training or laboratory infrastructure would enable diagnosis and treatment decisions to be made in the first patient encounter with potential for improved treatment outcomes (2).

Sputum is the primary specimen type for detecting pulmonary TB because it has a higher bacillary load than non-sputum specimens (4). However, up to one third of people being tested for pulmonary TB cannot expectorate sputum leading to a high burden of undiagnosed TB (5). In addition, sputum collection including sputum induction, if necessary, can be logistically challenging in some settings including infection control risks. In response to these limitations, alternative sample types such as tongue swabs are being evaluated. Tongue swab collection is non-invasive, readily adapted for self-collection, and has a very high sample collection yield ensuring that a sample is available even when sputum cannot be expectorated (6). The specificity of tongue swab specimens with molecular readouts, relative to sputum testing is consistently high. Sensitivity, however, is variable depending on population and methodology (7–13). Tongue swab specimen testing can be very sensitive among people with high bacillary loads, but less when applied to people with lower bacillary loads. Nonetheless, in patient populations and settings where sputum collection is not practical, tongue swab testing may offer a useful alternative (14).

The DASH® Rapid PCR System (Figure 1a) is a sample-to-answer platform that performs rapid, simple, durable, and low-cost infectious disease testing for multiple sample types such as respiratory swabs, whole blood, and plasma (15–17). We have adapted a previously published manual assay to detect *Mycobacterium tuberculosis* (MTB) that processes sputum via sequence specific capture to reduce amplification interferents that matched the sensitivity of culture (18, 19) for inclusion in the DASH test cartridge (Figure 1B). Since MTB is resistant to conventional bacterial lysis techniques, we also evaluated mechanical lysis and sonication as two pre-cartridge sample treatments for improved lysis.

**Figure 1.**
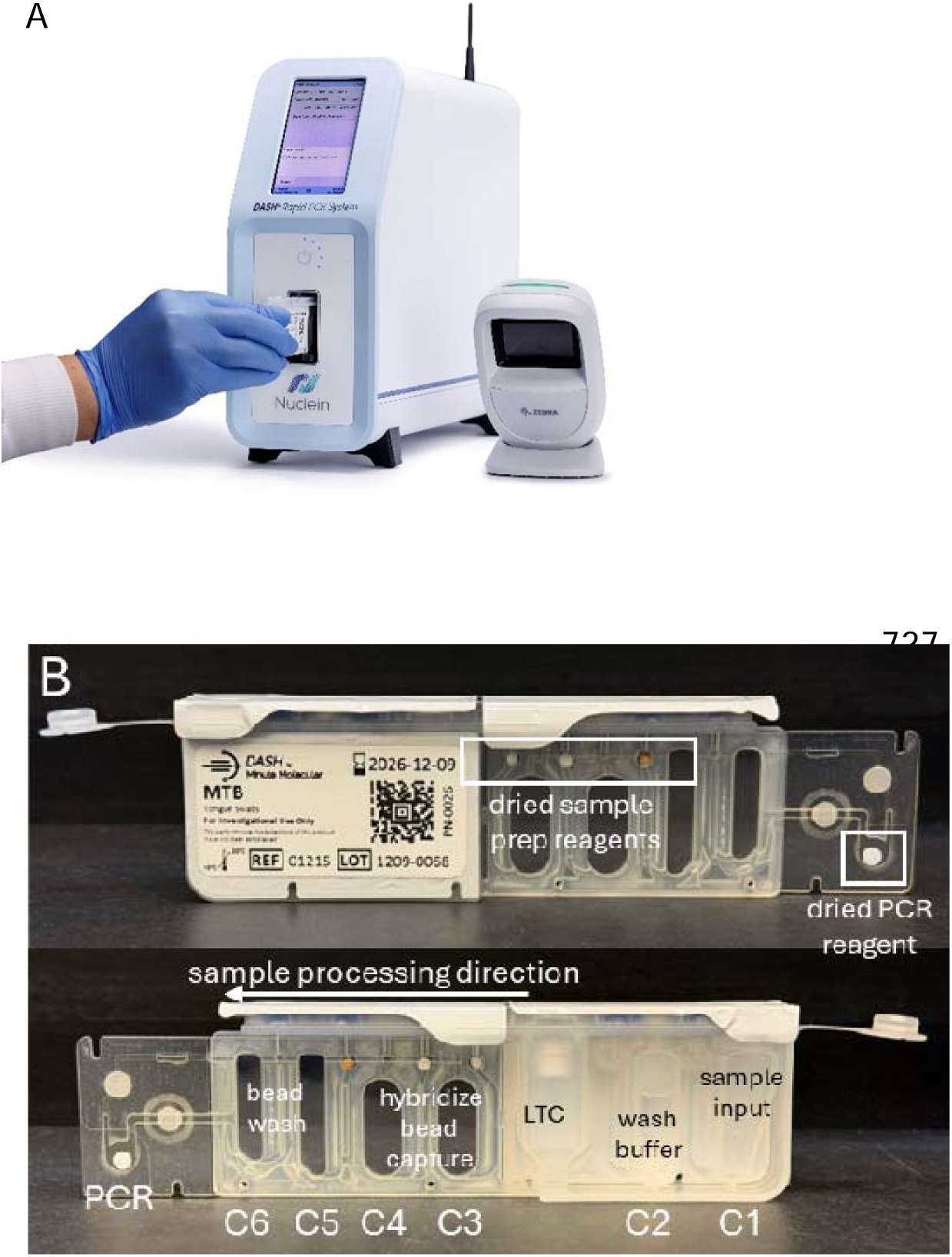
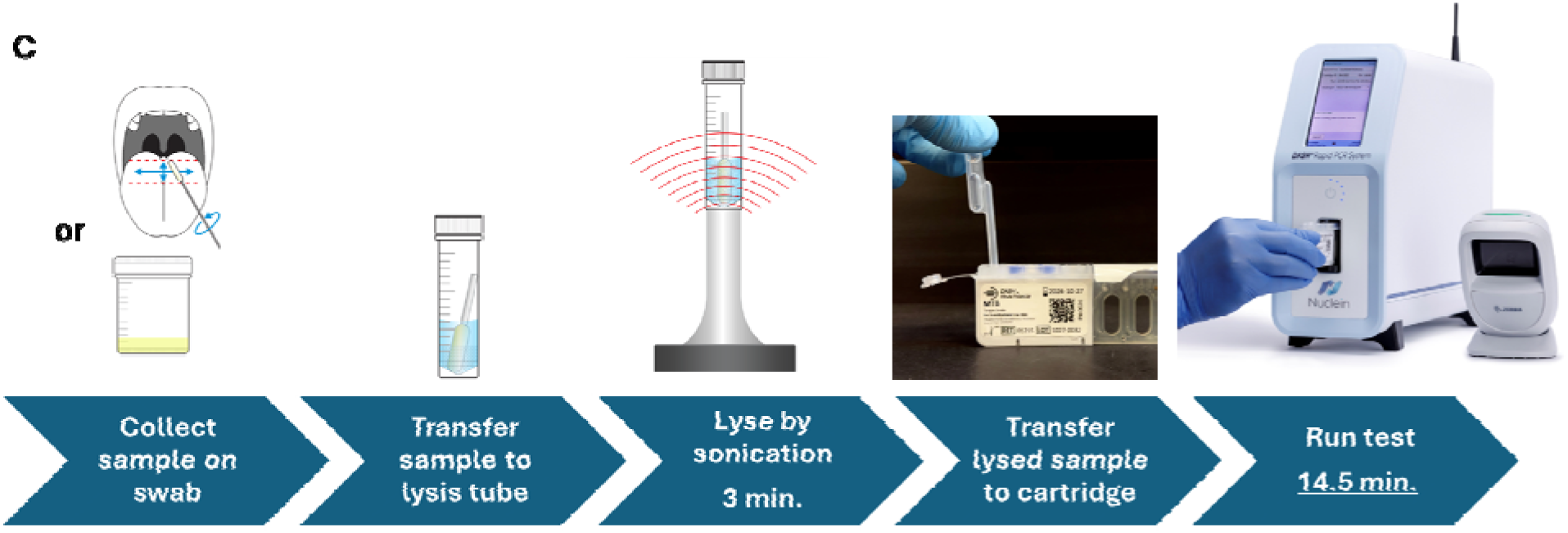
A) DASH® Rapid PCR System analyzer. B) RUO DASH®MTB cartridge contains all reagents required to run a test including sample prep and qPCR amplification and detection. The top image shows the front of the cartridge with stabilized reagents outlined in white boxes. The bottom image shows the back of the cartridge with the chambers labeled with the test functions. C1-C6 = Chambers 1-6; LTC = liquid transfer capsule. C) MTB assay workflow. Total time ≈ 20 minutes. 1) Specimens are collected from tongues or expectorated sputum using a flocked swab; 2) The swab containing the sample is placed in the lysis tube containing 600 µL lysis buffer and 150-200 mg 0.1 mm diameter glass beads; 3) The sample is sonicated for 3 minutes at power/amplitude of “4” with a downward force of 25-30N;4) 450 µL specimen is transferred to the sample chamber of the DASH MTB cartridge with a disposable pipet; 5) The cartridge is inserted into the DASH analyzer, and the MTB test is run automatically. After 14.5 minutes, an easy-to-read positive/negative result appears on the unit’s touch screen.

The objectives of this study were to optimize MTB assay performance on the DASH platform, determine the feasibility of testing tongue swabs or sputa as specimens by evaluating the test’s analytical and clinical performance, and demonstrate that the DASH analyzer is compatible with peripheral LMIC settings via external battery operation and cartridge stability at elevated temperatures.

## Materials and Methods

### Strains

MTB H37Ra was acquired from the American Type Culture Collection (ATCC; Manassas, VA) and used as a positive control for assay development. Table 1 contains strains used in the specificity studies. The concentration of *Mycobacterium tuberculosis* H37Ra cells from ATCC was estimated by comparing to quantified *Mycobacterium tuberculosis* H37Ra cells sourced from the University of Washington which were cultured and quantified by optical density as previously described (20). DNA from both cell sources was purified using Dynabeads® SILANE genomic DNA Kit (Thermo Fisher, Waltham, MA, USA) following manufacturer’s instructions. A standard curve was prepared by making tenfold dilutions ranging from 100,000 genomic copies to 10 genomic copies of the reference DNA. Ten-, 100-, and 1000-fold dilutions of the unknown DNA were prepared, and samples were evaluated using an assay targeting the IS*1081* insertion element (IS1081 F4, 5’- TGAACGCGCACTGACCA-3’, IS1081 R4, 5’-CTTTGGCCATGATCGACACTT-3’, IS1081 P2, 5’-TEX615/CTGCTACCTGCTGGGAGTATCCACT-3’).

**Table 1.** Summary of performance of organisms tested on the RUO DASH®MTB Test for cross-reactivity and microbial interference.

|  |  |  |  | Cross-reactivity | Microbial<br>interference |
| --- | --- | --- | --- | --- | --- |
| Organism | Source | Stock<br>conc.<br>(c/μL) | Conc.<br>tested<br>(c/rxn) | MTB<br>negative | MTB<br>positive |
| control (no potential<br>interferent) | n/a | n/a | n/a | 3/3 | 3/3 |
| *Mycobacterium*<br>*abscessus* | ATCC 19977D-5 | 3.74E+07 | 1.0E+06 | 3/3 | 3/3 |
| *Mycobacterium*<br>*avium* K-10 | ATCC BAA968D-5 | 3.08E+07 | 1.0E+06 | 3/3 | 3/3 |
| *Mycobacterium*<br>*genevense* | ATCC 51233 | 2.08E+06 | 1.0E+06 | 3/3 | 3/3 |
| *Mycobacterium*<br>*gordonae* | ATCC 35760D-5 | 2.21E+07 | 1.0E+06 | 3/3 | 3/3 |
| *Mycobacterium*<br>*intracellulare* | ATCC 13209 | 6.74E+06 | 1.0E+06 | 3/3 | 3/3 |
| *Mycobacterium*<br>*kansasii* | ATCC 12478 | 1.23E+07 | 1.0E+06 | 3/3 | 3/3 |
| *Mycobacterium*<br>*malmoense* | ATCC 29571 | 2.51E+06 | 1.0E+06 | 3/3 | 3/3 |
| *Mycobacterium*<br>*marinum* | ATCC BAA535D-5 | 1.74E+07 | 1.0E+06 | 3/3 | 3/3 |
| *Mycobacterium*<br>*smegmatis* | ATCC 23037D-5 | 3.59E+07 | 1.0E+06 | 3/3 | 3/3 |
| *Mycobacterium*<br>*terrae* | ATCC 15755 | 2.72E+06 | 1.0E+06 | 3/3 | 3/3 |
| *Bordetella pertussis* | ATCC 9797D-5 | 3.89E+07 | 1.0E+06 | 3/3 | 3/3 |
| *Psuedomonas*<br>*aeruginosa* | ATCC 47085D-5 | 2.05E+07 | 1.0E+06 | 3/3 | 3/3 |
| *Streptococcus*<br>*pyogenes* | ATCC 19615D-5 | 3.50E+07 | 1.0E+06 | 3/3 | 3/3 |
| *Streptococcus*<br>*salivarius* | BEI Resources,<br>NIAID, NIH as part<br>of the Human<br>Microbiome<br>Project: *Streptococ* *cus salivarius*,<br>Strain SK126, HM-121 | 1.55E+06 | 1.0E+06 | 3/3 | 3/3 |

### Swab control specimens

Swab used for contriving specimens were collected from healthy volunteers using Copan FLOQSwab (520CS01; Copan Italia, Brescia, IT) and stored at -80 °C until use.

### Swab clinical samples

Twenty-three pairs of swabs from TB positive donors provided by Rapid Research in Diagnostics Development for TB Network were used to compare lysis methods (Supplemental Methods; Table S1). The swab pairs were either Copan FLOQSwab (520CS01) or Steripack swab (60564RevC) stored dry at -80°C. Specimen data provided included sputum microbiological reference standard (MRS) results, sputum GeneXpert Ultra MTB/RIF (Xpert Ultra) semiquantitative results, and sputum culture results if the Xpert Ultra baseline result was negative. Five of the positive specimen pairs were High, 7 were Medium, 4 were Low, 5 were Very Low, 0 were Trace, and 2 were Xpert Ultra negative and culture positive.

Ten positive and 20 negative tongue swab specimens were provided by the Theron lab. Tongue swabs were collected using the Copan FLOQSwab (520CS01) from a cohort of adults self-reporting with presumptive pulmonary TB symptoms meeting WHO criteria (at least one symptom for people living with (PLHIV), ≥2 for those without HIV) and recruited at clinics in Cape Town, South Africa. The standard WHO-defined symptoms for TB were used (cough, weight loss, fever, drenching night sweats); with two required if living without HIV and one for PLHIV (for those without HIV, a cough needed to have persisted for at least two weeks; for PLHIV, any cough duration qualified). Tongue swabs were collected dry and stored at -80 °C following collection and until processed.

One hundred ten swab specimens (Copan FLOQSwab 520CS01) were previously collected from a cohort of South African participants by the South African Tuberculosis Vaccine Initiative as previously described (10, 13, 20). Participants with presumptive TB were enrolled in a clinical setting in Worcester South Africa between July 2021 and March 2023. Swabs not used in earlier studies were identified for testing in the present study and were shipped to Northwestern for processing from Seattle, Washington. All available swabs from participants with microbiologically confirmed pulmonary TB and available sputum Xpert Ultra semiquantitative data (N = 70) were selected for testing. An additional set (N = 40) of confirmed TB-negative participants was selected randomly for specificity calculations (STARD diagram). Samples were tested in a blinded fashion. Duplicate swab specimens from the same participants were tested in blinded fashion by using Xpert Ultra, as part of a previous study as described by Wood et al (13). All swabs were collected before sputum collection, or ≥1 hour after sputum collection.

### Sputum control specimens

Residual deidentified TB negative sputum specimens were provided by the microbiology department of the Mayo Clinic or were purchased from Tricore Reference Laboratories (Albuquerque, NM). Pooled sputum was prepared by vortexing twelve MTB-negative donor specimens totaling approximately 15 mL in a 50 mL conical tube with ∼5 grams of 5 mm acid-washed glass beads until homogenous.

### Sputum clinical specimens

One hundred sputum specimens were provided by Theron lab from a cohort described previously (21, 22). 51 were negative and 49 positive by sputum culture. Adults presenting with presumptive TB provided three sputum samples (each ≥1 mL, nonsalivary) over two consecutive workdays. The first sample underwent culture and smear microscopy, the second was tested with Xpert Ultra, and the third used for the DASH assay. GenoType MTBDRplus (v2.0; Bruker-Hain Life Sciences, Nehren, Germany) was performed on culture-positive isolates for MTBC and rifampicin resistance detection. (Figure 2).

**Figure 2.**
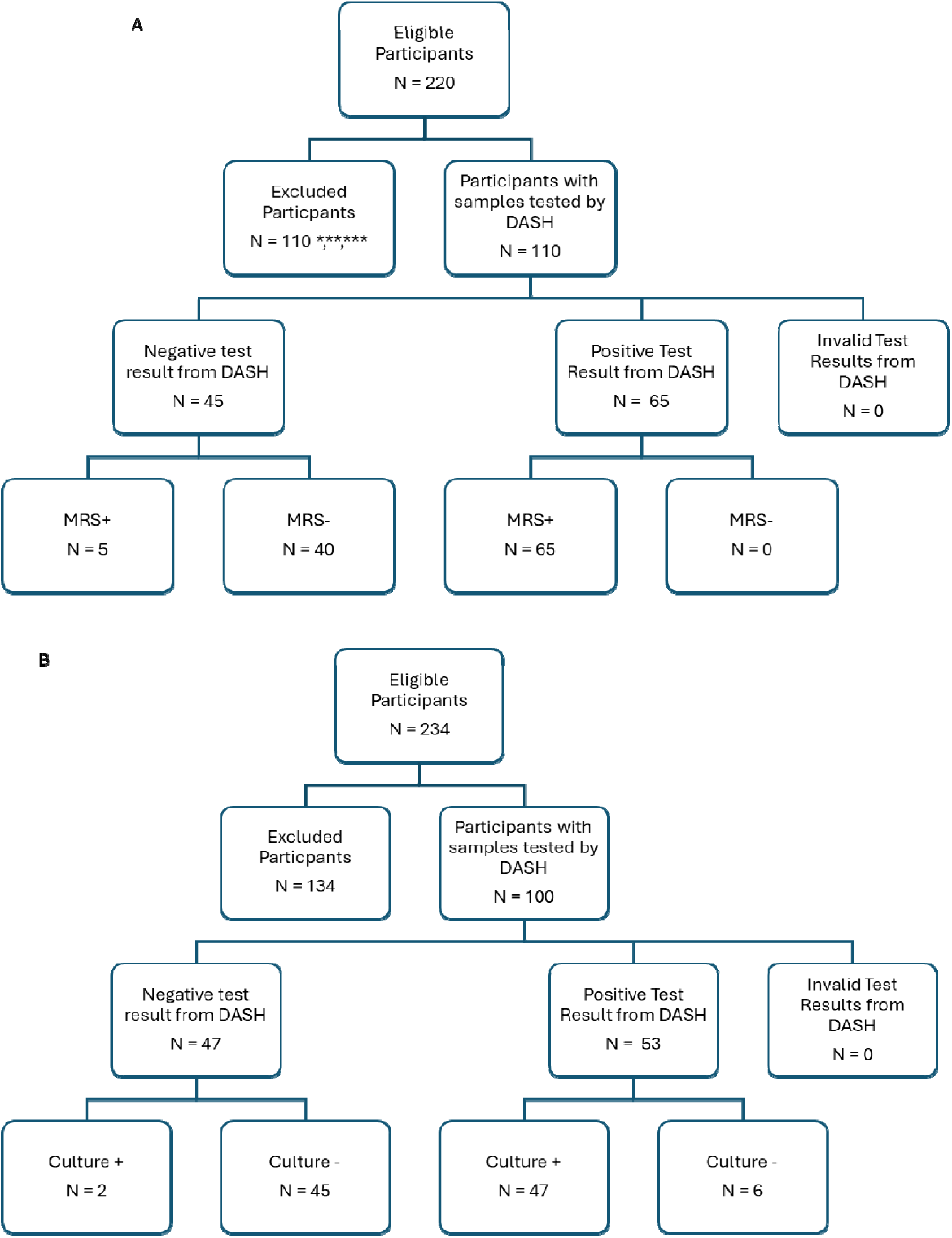
Study profiles for tongue swab and sputum specimen testing. STARDT (Standards for Reporting of Diagnostic Accuracy Studies) figures for tongue swab and sputum specimens. A) The STARD algorithm was conducted for inclusion of participants for the tongue swab specimen study. * Participants with confirmed negative TB were omitted randomly to test half of available n = 36; ** Participants with confirmed positive TB but no swabs were available for testing and/or used in previous studies n = 24;*** Participants with confirmed positive TB but no available Xpert Semiquantitative data available n = 50. B) The STARD algorithm was conducted for inclusion of participants for the sputum specimen study. 100 sputum specimens were randomly selected for DASH testing from prospectively recruited participants with valid culture and Ultra results.

### Sampling of sputum swabs

The volume of sputum retained on a swab was estimated as follows. Five swabs were dipped into aliquots of prepared pooled sputum and swirled 2-3 times, then were broken off into pre-massed 2.0 mL conical tubes. Three clean, broken-off swab heads were weighed and the average mass of the swab head plus tube were subtracted from the measured mass to calculate the mass of the sputum retained on the swab. This process was repeated with fifteen individual sputum specimens.

### DASH PCR System

The DASH platform achieved 510(k) FDA clearance in CLIA-waived settings for the DASH SARS-CoV-2 & Flu A/B multiplexed test and the DASH platform in 2024. Like central laboratory instruments, the DASH system extracts and concentrates target nucleic acid followed by qPCR amplification and detection. The fully integrated DASH cartridge (Figure 1B) consists of six sample-preparation chambers in the cartridge body, a liquid transfer capsule (LTC) which is used to transfer liquids between chambers, and a PCR chamber in the microfluidic extension. The analyzer extracts the target nucleic acid and binds targets of interest to paramagnetic particles (PMPs) via sequence-specific capture. The PMPs carrying the targets are separated magnetically from the lysis buffer, washed, and transported into the PCR chamber. The purified nucleic acid is eluted from the PMPs then amplified and detected by qPCR.

### Sonication Prototype

The sonicator consists of a circuit board (Sonics & Materials, Inc. Part No. KITVC544) connected to a probe (Sonics & Materials, Inc. Part No. CV301) and is powered by an AC-DC power supply (CUI, Inc. Model 3A-621DN24) outputting 24V at 2.5A. The sonication power/amplitude was modulated using a potentiometer (Honeywell Part No. 309NPC10K) wired into the board. To set the amplitude to reproducible values, a gauge was printed and adhered around the potentiometer such that the arc traced by turning the potentiometer begins at “0” and ends at “10”. The sonicator probe interfaces with the sample tube (USA Scientific 2 mL polypropylene screwcap tubes, #1420-9710; Ocala, FL, USA) by way of a custom 3D printed fixture that centers the bottom of the tube onto the probe.

### Research Use Only (RUO) DASH MTB Assay

The RUO DASH MTB Test is an automated qPCR assay intended for the qualitative detection of DNA from *Mycobacterium tuberculosis* complex organisms in tongue swab or sputum specimens. The test cartridge contains all the reagents required for the analyzer to automatically process a pre-lysed clinical sample and run the MTB test. Cartridge-specific reagents and cartridges were manufactured in the Northwestern University pilot manufacturing facility. Using DASH script software, the times, and temperatures of the sequence-specific hybridization, biotin-streptavidin binding, and the qPCR extension and denaturation steps were optimized.

The DASH MTB assay targets two multicopy regions of the MTB genome, IS*6110* and IS*1081,* which are detected in the same fluorescent channel. In lieu of primer sequences, the clarified Minimum Information for Publication of Quantitative Real-Time PCR Experiments (MIQE) guidelines allow publication of the reference sequence, anchor nucleotide (defined as a nucleotide located in the probe sequence), and amplicon length for each assay (23, 24). The IS*6110* amplicon context is NC_000962, 889,840, 100. The IS*1081* amplicon context is NC_000962, 1,169,640, 97. A synthetic DNA procedural control (PRC) is included to control for sample purification, nucleic acid amplification, and to monitor for the presence of inhibitors in the qPCR test. The synthetic DNA molecule contains a control amplicon and sequence-specific capture probe sequences used for capturing the IS*1081* DNA targets. The procedural control is fluorescently detected in a second channel on the DASH analyzer. At the completion of the test, the fluorescence readings from the MTB and the procedural control are processed by calculating the quantification cycle (Cq) of each channel and interpreting the result as either: MTB positive, MTB negative, invalid, or error.

### Test Workflow

The user collected either a tongue swab specimen or a sputum specimen on a swab (Figure 1C), inserted the swab into a flat-bottom 2 mL tube containing 600 µL of lysis buffer and 150-200 mg 0.1 mm diameter glass beads (Benchmark Scientific, Edison, NJ, USA). Samples were sonicated for 3 minutes at a power/amplitude of “4” with a downward force of 25-30N using a force gauge (SHIMPO; Model FG-3006; ELECTROMATIC Equipment Co., Lynbrook, NY, USA). Approximately 450 µL of specimen was transferred with a pipet to the DASH cartridge, the cartridge cap is closed, and the cartridge is inserted into the DASH analyzer. After 14.5 minutes, an easy-to-read positive/negative result appears on the unit’s touch screen.

### Limit of Detection

*Mycobacterium tuberculosis* H37Ra cells were diluted to 1e0 cells/µL and twofold serial dilutions were prepared three times in 10 mM Tris pH=8 (Invitrogen, Waltham, MA, USA), 15 % glycerol v/v (Acros Organics, now Thermo Fisher, Waltham, MA, USA), 0.05 % Tween-80 (Thermo Fisher). Twenty microliters of diluted MTB were added to a freshly thawed tongue swab, and the swab was processed using the sample workflow described above. Because 5/5 replicates were detected at 10 cells/swab, twenty replicates were collected at 2.5 cells/swab.

### Specificity

DNA from fourteen nontuberculous mycobacteria (NTM) or other pathogenic organisms that may be present in tongue swabs were tested for cross-reactivity and assay interference (Table 1). For the cross-reactivity evaluation, DNA or cells of each organism were added to a cartridge at the desired concentration with a tongue swab from an MTB-negative volunteer, in triplicate. For the interference evaluation, the cartridges were prepared as for cross-reactivity but with the addition of 60 MTB cells (3X LoD) per tongue swab.

Purified genomic DNA was used for all potentially interfering microbial organisms except for *Streptococcus salivarius*, where frozen bacteria were used. The concentrations of all nucleic acid additions were determined spectrophotometrically except for *S. salivarius* which was determined using a standard curve (supplemental methods).

### Clinical Evaluation of swabs

Tongue swabs were removed from -80 °C storage and placed directly into sonication tubes containing 600 µL lysis buffer and glass beads. Samples were left for a minimum of 5 minutes to equilibrate to room temperature before performing the test workflow.

### Amplification Efficiency with Sputum Specimens

*Mycobacterium tuberculosis* H37Ra cells were diluted to 2e5 cells/µL and five tenfold serial dilutions were prepared in 10 mM Tris pH=8 (Invitrogen), 15 % glycerol v/v (Acros), 0.05 % Tween-80 (Thermo). Each sample consisted of 10 µL of diluted MTB added to 100 µL negative pooled sputum, and each concentration was evaluated in replicates of 4. The entire prepared sample was collected on a flocked swab (Copan) and processed using the standard workflow.

### Clinical Evaluation of sputum specimens

49 culture positive and 51 culture negative sputum specimens were sampled by firmly pressing a Copan FLOQSwab into each sample tube while rotating the swab to load as much sputum as possible onto the swab head. The swab head was broken off into a flat-bottom sonication tube and processed via the test workflow.

### Cross-Contamination Studies

To confirm the absence of cross-contamination between assay runs, alternating runs of high positive and negative samples were run. The high positive sputum samples contained 14.3 million *Mycobacterium tuberculosis* H37Ra cells spiked into100 µL MTB negative pooled sputum (143,000 cells/µL), and the negative samples consisted of 100 µL negative pooled sputum. For both high positive and negative samples, 100 µL of pooled sputum was aliquoted to a 2 mL tube, the entire volume was sampled onto a swab and processed via the test workflow. The high positive samples were run on five analyzers, immediately followed by negative samples. This was repeated for a total of ten positive and ten negative samples.

### Cartridge Stability

One year stability at ambient temperature (22 °C) and 45 °C was established in real-time. Five replicates per time point of MTB positive (50 genome copies) and negative tests were performed at 1 month, 2 months, 4 months, 6 months, 9 months and one year. Stability was determined based on regression analysis of Cq values. Any percent change from time zero (baseline) was calculated between the test time point performance and the baseline performance [(T_test_-T_baseline_)/T_baseline_]*100 with 95% CI using the regression equation obtained for plotting the mean values. If the assay performance shift from the baseline to the current time point exceeds 10-15%, the cartridges were considered unusable.

### Statistics

Cq values were plotted against log cell number/swab to obtain a standard curve. Slope parameters and confidence intervals (CI) were estimated by linear regression of Cq values vs. log copy number (https://www.graphpad.com/quickcalcs/linear2/). The value of the slope was used to calculate PCR efficiency (25). Two-tailed Student’s t-tests and clinical sensitivity and specificity were calculated using Medcalc online package (https://www.medcalc.org/calc/diagnostic_test.php). Pearson correlation tests were used to generate R^2^ values to evaluate correlations between the two lysis procedures. Differences between the two tests were also examined graphically according to Bland & Altman (26). Plots were generated with the difference in Cq of the bead beating lysis method on the y axis and the Cq of heating lysis method on the x axis. The mean difference and the standard deviation of the differences were calculated, and lines were drawn corresponding to the mean and the mean ± 2 standard deviations. A t-test with the null hypothesis of a mean difference equal to zero (www.vassarstats.net/index.html) was performed. P values less than 0.05 were considered statistically significant. Analysis of Variance ANOVA calculator https://www.standarddeviationcalculator.io/anova-calculator was used to calculate the difference between two or more means or components through significance tests.

## RESULTS

### Comparison of lysis methods

The RUO DASH MTB Test is an automated qPCR assay intended for the qualitative detection of DNA from *Mycobacterium tuberculosis*. Since MTB is resistant to conventional bacterial lysis techniques, we initiated our studies by comparing mechanical lysis and sonication for sample processing prior to cartridge testing.

Paired oral swabs from 23 MTB-positive participants of varying bacterial loads were tested to compare nuclease inactivation/bead beating vs. sonication (Supplemental methods; Figure S1). Fourteen of the tongue swabs were positive in the sonication condition, and 11 were positive in the heating/bead beating condition (Table S1). The Cqs were compared for the 11 samples that were positive in both lysis conditions. Ten of the samples had lower Cqs (i.e., stronger signal) in the sonication condition, and one had a lower Cq in the heating/bead beating. This sample was considered an outlier by Bland Altman analysis (figure 2Sa). We hypothesize that there were significantly different bacterial loads between the paired swabs for this sample as has been reported in other studies (27, 28). When the apparent outlier was removed from the analysis, the two lysis methods are highly correlated (R^2^ = 0.95) (Figure 2Sb). The mean Cq difference between the sonication lysis method and the heat and bead beating lysis method was 1.2 (95% CI: 0.66, 1.71; (p=0.0006). Sonication was thereby deemed to be the more effective lysing treatment and was used as the lysis step in all subsequent testing.

### Analytical limit of Detection

The limit of detection (LOD) of the DASH MTB test was determined using negative tongue swab samples spiked with MTB H37Ra (ATCC). Five of five swabs contrived with 10 cells/swab were detected, and 20 of 20 contrived with 2.5 cells/swab were detected. Lower dilutions were not tested because the sampling error at 1 cell/swab was too great, and the LOD was declared to be 2.5 cells/swab.

### Specificity

#### Cross-Reactivity

DNA from fourteen NTM strains and/or other bacterial strains that are found in the mouth were tested to determine if they cross reacted in the DASH® MTB test cartridge (Table 1). No positive results were observed in the MTB channel. The procedural control average Cq for the tongue swab control was 29.6±0.3 and the average Cq for the potentially cross-reactive species was 29.7±0.6 demonstrating that there is no real difference between the means (P=0.7779). Thus, a valid, negative result was obtained for all replicates indicating a lack of cross-reactivity at the concentration tested.

#### Microbial Interference

The same 14 bacterial strains used for cross-reactivity assessment were used to contrive MTB positive samples with 60 MTB cells/swab, in triplicate. A valid, positive result was obtained for all replicates tested, indicating a lack of assay interference (Table 1). The average Cq of the positive controls without addition of any potentially interfering strain was 31.4± 0.7, and the average Cq of the samples spiked with potentially interfering strains was 30.9±0.9, demonstrating that there is no real difference between the means (P=0.35) and indicating that the assay performance was similar with and without the presence of the potentially interfering strains.

### Swab Specimen Testing

A pilot study of 30 swab specimens (10 positive, 20 negative) was performed using the DASH assay with no invalid results. The DASH assay had 70% sensitivity and 95% specificity compared to sputum culture. The Xpert Ultra test applied to tongue swabs from the same participants yielded a 67% sensitivity and 90% specificity (Table 2). The pilot study results indicate that tongue swab specimens are compatible with the DASH MTB assay when combined with the sonication lysis step.

**Table 2.** Tongue swab specimen pilot study.

|  | Sputum |  | Tongue Swab |  |
| --- | --- | --- | --- | --- |
| TB Status | Culture | Ultra | Ultra | DASH |
| Positive | 10/10 (100%) | 10/10 (100%) | 6/9 (67%) | 7/10 (70%) |
| Negative | 20/20 (100%) | 20/20 (100%) | 9/10 (90%) | 19/20 (95%) |

A second larger study of 110 blinded swab specimens (70 positive, 40 negative) (Table S2 and S3) was performed using the DASH assay with no invalid results. The DASH assay was 91% sensitive and 100% specific when compared to the microbiological reference standard (MRS) (Table 3). When compared to the Xpert Ultra sputum test results, the DASH assay accurately detected the High and Medium semiquantitative (94% and 100%) samples and more moderately detected the Low and Very Low (88% and 80%) samples. The DASH and the Xpert Ultra tongue swab results had nearly equivalent sensitivity (91% vs. 90%) and equivalent specificity (100%).

**Table 3.**
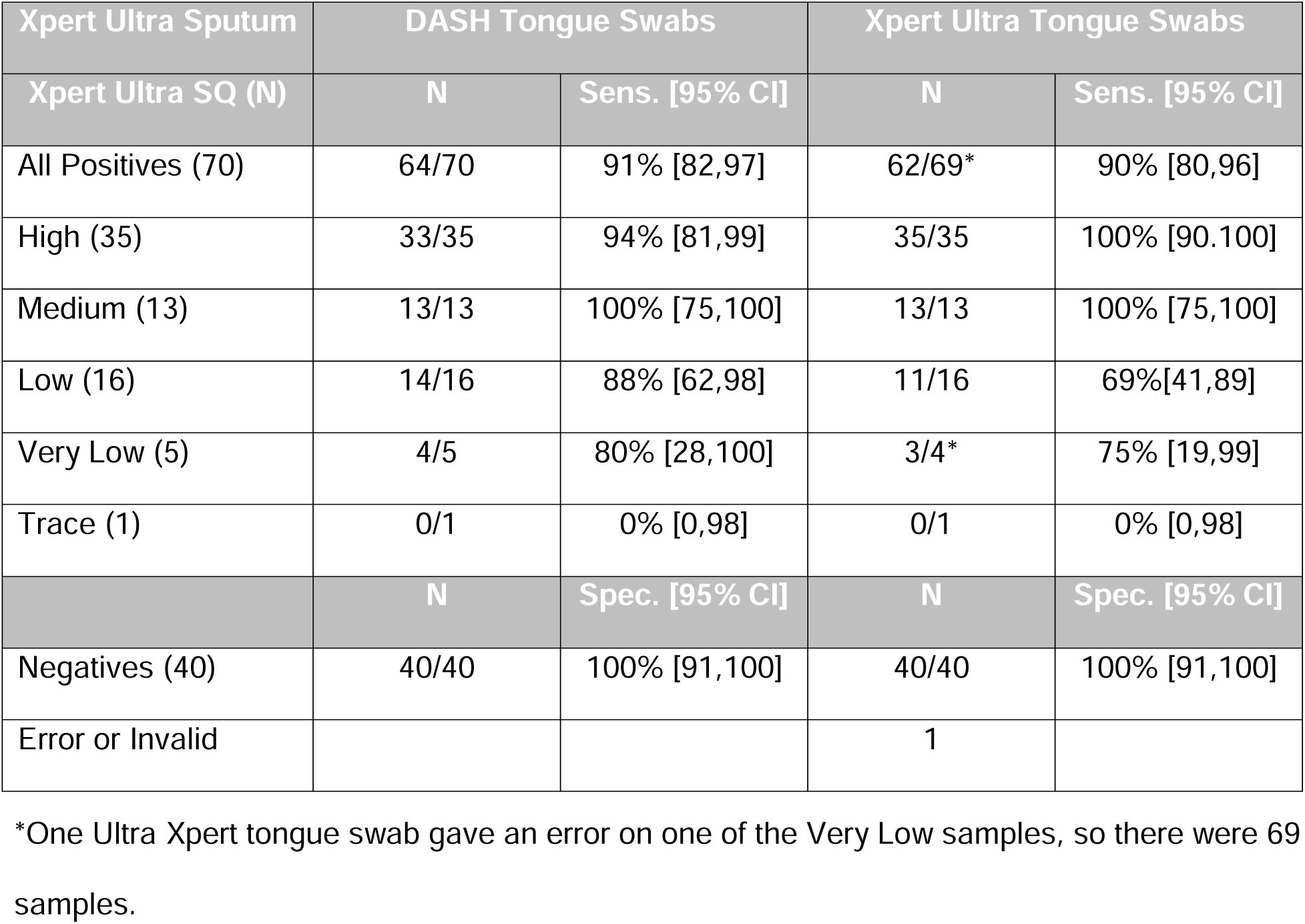
One-hundred and ten tongue swabs were tested in DASH MTB assay with results compared to Ultra MTB test.

#### Sputum testing

Sampling sputum with a swab was first reported by Steadman, et al. in a multi-country diagnostic accuracy evaluation of late prototype versions of two nPOC MTB tests (12). Sputum usually has a higher bacillary load than non-sputum specimens in pulmonary TB (4). Therefore, we performed a series of studies to determine if sputum can be adequately processed in the DASH platform using the tongue swab workflow described above.

### Characterization of Sputum as a Specimen

The challenges of isolating amplification-ready nucleic acid from sputum include the high viscosity and heterogeneity of the sputum matrix, high concentrations of amplification inhibitors, uneven distribution of the bacilli, relatively small amounts of MTB nucleic acid compared to human nucleic acid, and the bacilli’s resistance to lysis due to its thick cell wall with waxy coat (19). A benefit of the sonication pretreatment is that the sample is lysed and thoroughly homogenized before being transferred to the DASH cartridge for nucleic acid extraction, amplification, and detection.

To determine the volume of sputum sampled by the Copan swab, we prepared a pooled sputum specimen, dipped a swab into the specimen 5 times and weighed the swab head to determine the average volume collected to be 95.0±19.3 µL (range 80 to 127 µL). We then tested 15 individual sputum specimens and observed that the average volume collected was 136.2±62.5 µL (range 65 to 260 µL) which was slightly more than with the pooled sputum. Moreover, the quantity collected was more variable. To test if the large range of sputum volume would be processed efficiently by the DASH platform, we added zero (N=3), 50 (N=5), and 300 (N=5) µL of pooled sputum to the reactions spiked with MTB. The mean MTB Cqs of the 3 sputum volumes were 30.5±0.3, 30.5±0.4, and 30.5±0.9, respectively. There was no significant difference between the means (p=0.98809) indicating that the amount of sputum sampled by the swab will not interfere with the sample processing in the DASH cartridge.

### Assay Efficiency in the Presence of Sputum

To determine if using sputum as the specimen interferes with the PCR efficiency of the MTB assay, the test linearity was determined by examining the relationship between Cq and MTB concentration. The DASH MTB assay was linear (R2= 0.99) from 20 to 2 million cell spiked sputum specimens (Figure 3). The assay correctly identified MTB in all spiked samples. The PCR efficiency of 97.4% was calculated from the line’s slope of -3.39 [95%CI slope: -3.57,-3.21; p < 0.001]. A highly efficient qPCR assay is required for optimal product yield, and thus low LOD, and indicates that the sputum did not inhibit the assay (29).

**Figure 3.**
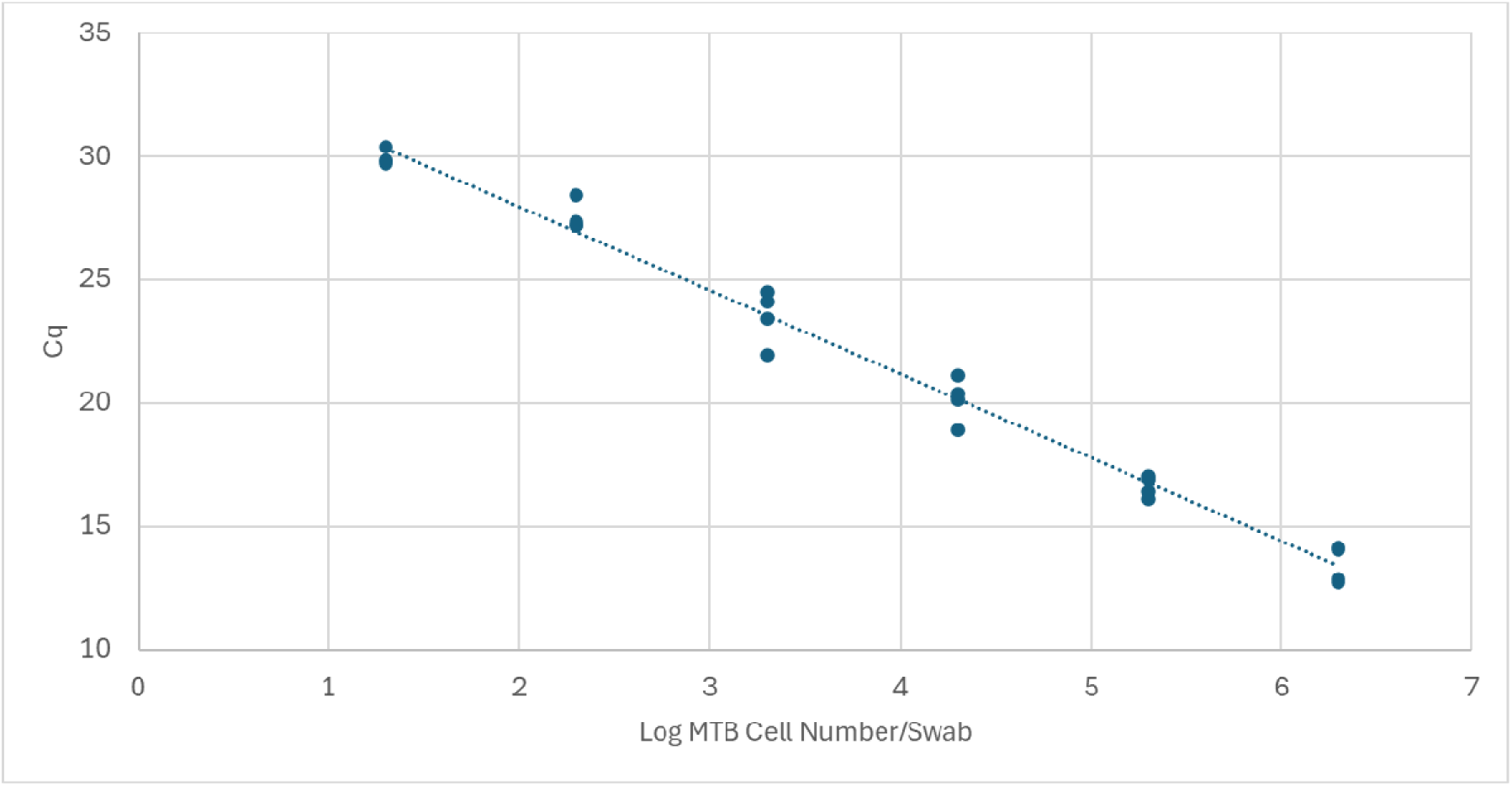
Linear and Efficient MTB amplification in the presence of sputum indicates a lack of PCR inhibition caused by sputum. Standard curve of log copies *Mycobacterium tuberculosis* (x) versus Cq (y). Equation of line: y = -3.39X + 34.74 [95%CI slope -3.57,-3.21]; R2 = 0.99. PCR efficiency calculated from the slope 97.4%.

### Sputum Clinical Testing

One hundred sputum specimens (Table S4 and S5) were tested by the DASH assay with no invalid results. Overall sensitivity was 96% and specificity was 88% relative to culture results (Figure 4). The Xpert Ultra sensitivity and specificity for these specimens were identical to DASH. DASH had 100% agreement with the Xpert Ultra semiquantitative categories of High, Medium, Low, and Very Low and 57% agreement with Trace samples. Overall, nine samples had false-positive results in either the DASH or Xpert Ultra test: three were positive in both assays, and three were positive in one assay or the other (Figure 3B). Of these 9, 5 had prior TB disease episodes, and the 6 Xpert Ultra-positive samples had Trace semiquantitative results.

**Figure 4.**
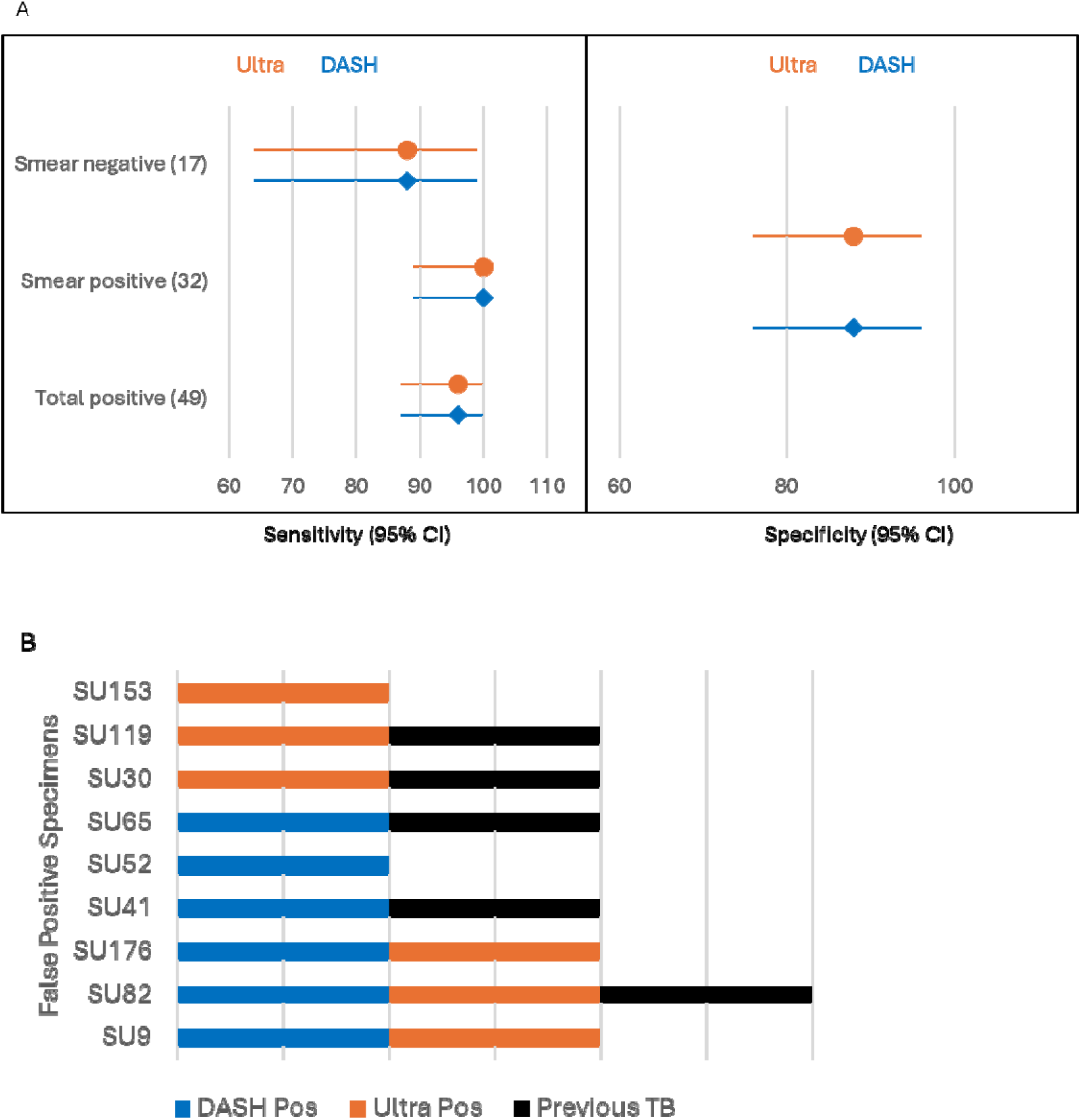
A) Forest plot of Xpert Ultra (orange circles) and DASH (blue diamonds) sensitivity and specificity on sputum specimens with culture as the comparator. Both assays had identical positive and negative agreement percentages to culture. B) Nine samples were false positive in either the DASH and/or Xpert Ultra assays, and 5 of the participants reported previous TB disease. A bar is present if the sample was positive with Xpert Ultra (orange), DASH (blue), or the participant reported previous TB disease (black).

#### Near Point-of-Care Compatibility

To be compatible with LMIC peripheral healthcare settings, the DASH MTB test must function with low-quality or intermittent power via external battery operation. It also needs to use tightly sealed cartridges that retain amplicons generated during the testing (thus eliminating the need for separate pre- and post-amplification rooms required for laboratory-based nucleic acid amplification tests), and to have long term cartridge stability in settings that lack refrigeration and air conditioning.

### Lack of Cross-Contamination

A study was conducted to demonstrate that cross-contamination does not occur when processing specimens with the sonication step followed by the single-use, self-contained DASH® MTB test cartridges. High positive samples were run in five DASH® analyzers. Negative samples were processed in the same DASH analyzer immediately following the positive specimen. This testing scheme was performed twice for a total of twenty runs in five analyzers, resulting in 10 positive and 10 negative correctly reported specimens with no cross contamination observed.

### Cartridge stability

The effect of long-term storage at ambient (22 °C) and elevated (45 °C) temperatures for over one year was determined. No difference in performance after 373 days of incubation compared to day zero was observed with cartridges incubated at either ambient or 45 °C (Figure S3).

### Battery operation

An Anker Portable Power Station (PowerHouse 256Wh; Model A1720) was used to operate a DASH analyzer using a custom 12V car charger adapter to a 4-pin DIN connector for 33 back-to-back 15-minute tests totaling approximately 8.25 hours of operating time without any error.

## Discussion

There is an urgent need for TB tests to be located at peripheral healthcare centers for rapid and accurate MTB detection enabling providers to initiate therapy in the same clinical encounter (30). In this study, we demonstrated the high analytical and clinical sensitivity and specificity of the DASH MTB assay combined with a pre-cartridge sonication step using either tongue swabs or sputum specimens.

Tongue swabs are recognized as a promising alternative diagnostic specimen to sputum with high specificity but generally lower sensitivity, with an observed correlation between sputum bacterial loads as measured by Xpert Ultra semiquantitative results and tongue swab positivity (13). Since 2015, well over 21 studies have explored the use of tongue swabs as specimens (31). From these studies, 4 key assay steps have been identified to improve test sensitivity: nuclease inactivation; optimal bacterial lysis; maximizing the specimen volume tested either via multiple PCR runs, high-volume PCR runs or solid phase extraction/concentration; and multicopy target amplification (10, 27) . The DASH test was designed with these key steps in mind. The combination of the DASH lysis buffer and sonication inactivates nucleases present in oral specimens and lyses the notoriously durable MTB cells. The specimen is then transferred to the DASH cartridge where the quantity of MTB target analyzed is maximized via sequence-specific capture. The multicopy targets IS*6110* and IS*1081* are amplified and detected yielding the high sensitivity (91%) and specificity (100%) observed in the study of 110 banked tongue swab specimens.

Sputum specimens, when available, are the preferred MTB specimen, as they are predicted to have the lowest false negative rate due to higher bacillary loads (32). The challenges of isolating amplification-ready nucleic acid from sputum such as high sample viscosity and high concentrations of PCR inhibitors are addressed in the DASH assay by the pre-cartridge sonication step and the sequence specific DNA extraction method used in the DASH cartridge (10, 19). In this study, we determined the range of sputum volume sampled when the Copan swab was swirled in sputum and demonstrated that this volume did not affect MTB detection or reduce PCR efficiency (Figure 3).

The DASH MTB test and the Xpert Ultra test had identical sensitivity (96%) and specificity (88%) with this sample set of 100 sputum specimens (Figure 4A). Nine samples were false positive for one or both assays (Figure 4B) which may imply that these specimens were near the LOD of both assays. The semiquantitative values for the six false positive specimens in the Xpert Ultra test were Trace positive. A proportion of Trace positives have been attributed to residual MTB DNA from previous TB disease (33–36), and five of the nine false positive specimens from this study were collected from participants reported to have had previous TB disease. Recently, in high burden TB settings, it has been reported that up to one half of patients with sputum Trace results were diagnosed with TB disease within 12 months (37–39) which indicates that a Trace result may reflect paucibacillary disease when interpreted within the appropriate clinical and imaging framework rather than an indeterminate result.

WHO has recently published updated target product profiles (TPP) for tuberculosis screening tests (40) with three types of tests defined on the basis of healthcare setting, the complexity of the test, the infrastructure, and the equipment and skills required: 1. Point-of-care tests defined as instrument-free and not requiring any laboratory infrastructure (example: lateral flow test); 2. Near point-of-care tests defined as preferably battery operated instruments that do not require laboratory infrastructure (example: portable nucleic acid amplification test platform); and 3. Low complexity tests requiring an instrument and basic laboratory infrastructure with basic technical skills to use it (example: GeneXpert 10 color platform) (30). We have demonstrated that DASH is compatible with nPOC settings in that DASH MTB cartridges are stable at elevated temperatures, the DASH analyzer can be powered by an external battery, and the lack of cross contamination eliminates the need for separate pre- and post-amplification rooms. In our performance testing, we demonstrated that the DASH MTB assay met or exceeded the WHO TPP minimal sensitivity requirements for both sputum (85%, 90%) and non-sputum i.e., tongue swab specimens (75%, 80%), for both nPOC and low complexity settings, respectively. The specificity of the sputum testing was less than the >95% optimal, but the non-sputum specificity of 100% exceeded the optimal value.

There are limitations to this study. The procedural control is a synthetic DNA molecule which does not report on the efficiency of cell lysis. In our next assay design iteration, the synthetic construct will be replaced by a genetic target from *Bacillus atrophaeus* spores which will confirm that efficient lysis has occurred. Additionally, the pre-DASH cartridge sonication step uses a prototype sonicator that is not ready for transfer to untrained operators. The next generation sonicator will be portable and battery operated and will not require training to set the applied pressure to the tubes during sonication. The swab and sputum specimens had all been previously frozen, and the freeze/thaw may have overestimated or underestimated the lysis efficiency which could have skewed the test sensitivity. Moreover, the clinical sputum and tongue swab specimens were tested by well-trained laboratorians. Future evaluations in nPOC settings with nonlaboratory users processing fresh tongue swab and/or sputum specimens are justified to demonstrate that laboratory training is not required and that the test performance is comparable to that with frozen samples. Finally, the specimens tested were selected randomly from storage not from consecutively collected specimens, which may have inadvertently introduced sampling bias.

Despite these limitations, the initial characterization of the DASH MTB test demonstrated high clinical and analytical sensitivity and specificity, robustness against cross-contamination, and compatibility with settings that lack air conditioning and consistent power. Sensitive detection of MTB from tongue swabs and sputum plus the rapid time to result indicate high potential for use with same day diagnosis and treatment of active tuberculosis infection. A further advantage of the DASH platform is that in addition to detecting MTB, it can be used for multi-disease detection from a range of sample types such as nasal swabs for a multiplexed SARS-CoV-2, Influenza A, and Influenza B test (17) to whole blood or plasma for an active hepatitis C diagnostic test (16) thus optimizing resources and expanding access to testing outside siloed testing programs (2).

## Supporting information

Supplemental Methods and Results

## Data Availability

All data produced in the present work are contained in the manuscript and online at McFall, Sally (2026), Development of a Rapid Automated Point-of-Care Test for Mycobacterium tuberculosis Detection from Tongue Swabs and Sputum Specimens on the DASH Rapid PCR System, Mendeley Data, V1, doi: 10.17632/mybvt96x97.1

## Acknowledgement

The authors thank past and present members of the Center for Innovation in Global Health Technologies, the Clinical Mycobacteriology & Epidemiology (CLIME) research group, and study participants. We thank Zelalem Temesgen, MD for the gift of residual deidentified TB-negative sputum specimens and the Rapid Research for Diagnostics Development in TB Network (R2D2) funded by the National Institute of Allergy and Infectious Diseases of the National Institutes of Health (award U01AI152087) for providing us with paired tongue swab specimens for the lysis study. We thank Nuclein LLC for providing molded cartridge bodies that were used to assemble DASH® MTB cartridges at Northwestern University.

