## Supplemental Methods and Results for "Development of a Rapid Automated Point-of-Care Test for *Mycobacterium tuberculosis* Detection from Tongue Swabs and Sputum Specimens on the DASH® Rapid PCR System"

Supplemental Information

Methods

*Quantification of S. salivarius*

For *S. salivarius* quantification, nucleic acids were purified using Dynabeads Silane Viral NA kit following manufacturer’s instructions (Thermo Fisher), and the concentration was estimated using a universal bacterial16S qPCR assay (338 F, 5’-ACTCCTACGGGAGGCAGCA-3’ and 518 R, 5’- ATTACCGCGGCTGCTGG-3’) (1) and a standard curve of known amounts of *Mycobacterium tuberculosis* DNA, which contains a single copy of the 16S rRNA gene.

*Lysis Study*

Twenty-three pairs of swabs provided by Rapid Research in Diagnostics Development for TB Network collected from the same patient were tested to compare MTB lysis by bead beating or sonication. The swab pairs were either from Copan FLOQSWAB (520CS01) or Steripack swab (60564RevC). Steadman et al., (2) demonstrated that there was no statistically significant difference between the amount of MTB detected from these different swabs. Post lysis procedures, MTB DNA was extracted from the samples using manual sequence-specific capture and qPCR in the 5-plex Qiagen (Hilden, Germany) Rotor-Gene Q thermocycler (Figure S1).

Bead beating

The following method was based on a method developed by Steadman et al. for processing tongue swabs. (2). Clinical tongue swabs (swab head down) were transferred to 1.5 mL conical screw cap tubes containing 500 µL TE (10mM Tris pH 8.0, 1mM EDTA pH 8.0). The samples were vigorously vortexed for 15 seconds then incubated for 10 minutes in a Benchmark Multi-Therm heater-shaker (Benchmark Scientific, Edison, NJ, USA) set to 100 °C without shaking. After incubation, the samples were once again vortexed for 15 seconds. Next, the entire volume (400-450 µL) from each tube was transferred to BeadBug™ tubes (2 mL capacity, Millipore Sigma Z763837) containing 150-200mg of 0.1 mm acid washed Silica glass beads (Benchmark Scientific D1131-01). Samples were bead beaten in a BeadBug™ benchtop homogenizer (Benchmark Scientific model D1030) for three 1-minute rounds at 3000 rpm with 1-minute breaks after each of the first two rounds. After processing, 300 µL were taken from each tube and transferred to 1.5 mL conical tubes containing enough dried Sodium Dodecyl Sulfate (SDS) and Tris pH 8.0 to bring the final concentrations to 1% v/v and 30 mM, respectively. Tubes were vortexed for 20 seconds to resuspend the dried lysis buffer components.

**Sonication**

Clinical tongue swabs were transferred to 2 mL screw cap tubes (swab head down) containing 150-200 mg of 0.1 mm silica glass beads and 500 µL Lysis Buffer. Samples were sonicated for 80 seconds at a power/amplitude of “3.5”. Downward pressure applied to the sample tube during sonication was determined empirically by the operator. The operator applied the minimum force necessary to cause the glass beads to visibly jostle around, and to prevent melting through the tube the force was reduced as the tube bottom softened and better contact was made between the tube and probe.

*Manual DNA extraction*

MTB DNA was manually extracted from the lysed specimens using our published method with a few modifications (3). A lyophilized pellet containing active ingredients of 0.5 mg Proteinase K (Gold Biotechnology, St. Louis, MS, USA) 6.2 µg CaCl_2_, 5 pmol biotin-labeled capture oligonucleotides targeting IS6110 and IS1081 (IDT, Coralville, IA, USA), and 500 copies of a custom gBlock™ process control (IDT) was added to each tube and vortexed for 20 seconds for resuspension. Capture probes contained a 5’ dual-biotin moiety, included a spacer of 5 adenine residues prior to specific sequences, and were HPLC-purified. Samples were incubated in a heater-shaker for 2 minutes at 55 °C at 1500 rpm then transferred to a separate preheated heater-shaker for 2 minutes at 100 °C at 1500 rpm. The samples were cooled in a room temperature aluminum block for 2 minutes. 30 µL of a solution of 3M NaCl and 0.8M MgCl_2_ were added to each tube and mixed in with brief vortexing. The samples were placed into a 68°C heater-shaker at 1500 rpm for 5 minutes then cooled in a room temperature aluminum block for 2 minutes. Dynabeads M-270 Streptavidin paramagnetic particles (PMPs) (Life Technologies) sufficient for 300 µg per sample (6 µL of a 50 mg/mL suspension) were washed two times in 0.5 mL TT buffer (10mM Tris pH 8.0, 0.01% Tween-20) and resuspended in their initial volume of TT buffer. 6 µL of washed PMPs were added to each sample tube and briefly vortexed to mix. Samples were incubated at 68°C at 1500 rpm for 5 minutes then cooled in a room temperature aluminum block for 2 minutes. Samples were centrifuged for 1 second to collect the samples and placed on a DynaMag ™-2 magnetic stand (Invitrogen, 12321D) where PMPs were collected, and the supernatant was discarded. PMPs were washed in 0.5 mL TT and transferred to a clean tube. PMPs were collected and washed again in 0.5 mL TT, for a total of two washes. All remaining solution was carefully removed with a pipet. Finally, 11.5 µL of TT was used to resuspend the PMPs in each tube. Samples were eluted at 75 °C for 3 minutes with 1500 rpm shaking. PMPs were pelleted on a magnetic stand, and eluted DNA was transferred to a clean tube. Tubes were placed in a -80 °C for later qPCR amplification.

*PCR Amplification*

PCR mastermix for a 25 µL reaction volume (20 µL master mix and 5 µL of eluted DNA) consisted of: 0.2 mg/mL bovine serum albumin (BSA; Thermo Fisher Scientific Inc.; Waltham, MA), 0.2% Tween-20 (Thermo Fisher), 150mM trehalose (Life Sciences Advanced Technologies Inc.; St. Petersburg, FL), 10% glycerol (Acros Organics, Now Thermo Scientific Chemicals), 62.5mM bicine pH 8 (Sigma-Aldrich; St. Louis, MO ), 135mM potassium glutamate pH 7.5 (Sigma-Aldrich), 2 mM magnesium chloride (Sigma-Aldrich), 0.325 each dNTP (Life Technologies Corporation; Grand Island, NY), 3.75 U RMS Hawk Fast Z05 polymerase (Roche Diagnostics Corporation; Indianapolis, IN), and oligos targeting the MTB insertion sequences IS*6110* and IS*1081* as used in Steadman et al 2024 (2). The IS*1081* primers also amplify the gBlock™ process control. All oligos were present at 200 nM. Amplification was performed in a 5-plex Qiagen (Hilden, German) Rotor-Gene Q thermocycler. Cycling conditions were as follows: 95°C initial DNA melt for 2 minutes followed by 50 cycles of 95 °C 15-second melt and 60 °C 45-second extension.

Supplemental Tables

Table S2. Patient characteristics of tongue swab donors.

| Demographics (N=110) | Positives (N=70) | Controls (N=40) |
| --- | --- | --- |
| Average Age (Range) | 36 (18-68) | 40 (19-67) |
| Male Sex (%) | 46 (66) | 19 (48) |
| Ethnicity | | |
| Asian | 0 | 1 |
| Black | 19 | 10 |
| White | 1 | 0 |
| Mixed Race | 50 | 29 |
| HIV Status | | |
| HIV Positive (%) | 18 (26) | 11 (28) |
| HIV Negative (%) | 52 (74) | 29 (72) |
| Bacterial Confirmation of TB from Sputum Testing | | |
| MRS Positive | 70 | 0 |
| MRS Negative | 0 | 40 |
| Culture Positive | 67 | 0 |
| Culture Negative | 0 | 43 |
| Xpert Ultra Positive | 70 | 0 |
| Xpert Ultra Negative | 0 | 40 |

MRS=Microbial Reference Standard.

Table S4. Patient Characteristics of sputum specimen donors.

| Demographics (N=100) | Positive (N=49) | Negative (N=51) |
| --- | --- | --- |
| Age Average (Range) | 36 (18-72) | 40 (19-62) |
| Male Sex (%) | 25 (51) | 28 (55) |
| BMI | | |
| <18.5 (underweight %) | 34 (69) | 16 (31) |
| 18.5-24.9 (healthy weight %) | 12 (24) | 24 (47) |
| 25-29.9 (overweight %) | 2 (4) | 5 (10) |
| >30 (obese %) | 1 (2) | 6 (12) |
| HIV Related Information | |  |
| HIV-Positive (%) | 16 (33) | 8 (16) |
| HIV-Negative (%) | 33 (67) | 43 (84) |
| TB History | | |
| Previously Diagnosed with TB (%) | 25 (51) | 21 (41) |
| Not Previously Diagnosed with TB (%) | 24 (49) | 30 (59) |
| Bacteriological Confirmation of TB (sputum 1) | | |
| Smear Positive/Culture Positive n (%) | 32 (65) | 0 |
| Smear Negative/Culture Positive n (%) | 17 (35) | 0 |
| Smear/Positive/Culture Negative n (%) | 0 | 1 (2) |
| Smear Negative/Culture Negative n (%) | 0 | 50 (98) |
| Xpert MTB/RIF Ultra (sputum 1 semiquantitative result) n | 53 | 0 |

Supplemental Figure Legends

Figure S1. A) Workflow for heating and bead beating. The swab and TE buffer are heated to 95°C for 10 minutes to inactivate nucleases; the supernatant is transferred to a bead beating tube and subjected to 3 rounds of bead beating for 1 minutes; and 300 µL of the lysed sample is added to a tube containing dried specific capture lysis buffer. B) Workflow for sonication lysis. The swab in the specific capture lysis buffer is subjected to sonication for 80 seconds; the supernatant is transferred to a clean tube. C) Workflow for specific capture and qPCR amplification and detection. 300 µL of lysate is treated with proteinase K (PK); the sample is then heated to 100°C to denature the residual PK and melt the double stranded DNA; salt is added to the tube, and the biotin-labeled sequence specific capture probes hybridize to the target DNA; streptavidin coated paramagnetic particles (PMPs) are added to the tube to bind to the biotin-probe/MTB DNA target; the PMPs are washed to remove potential PCR inhibitors, and the target DNA is eluted from the PMPs at 75°C for 3 minutes; and the eluate is added to qPCR reactions targeting MTB IS*6110* and IS*1081.*

Figure S2. A. Bland-Altman analysis. Mean Quantification Cycle (Cq) values of the two lysing methods (x) versus the difference between the two methods (y). Dotted blue line = mean difference between methods (-0.94). Dotted red lines = ± 2 standard deviations of mean difference (1.18, -3.05). The potential outlier point is circled. B. Cq Correlation between sonication (x) and bead beating (y) lysis methods with 1 outlier data point removed. Equation of line y = 0.9X + 3.4 [slope 95% CI: 0.72, 1.1; p <0.001] R^2^ = 0.94.

Figure S3. Stability of DASH MTB Cartridge after one year incubation at 22°C (ambient) and 45°C. Five replicates per time point of MTB positive (A & C) and the PRC for MTB negative (B & D) tests were performed at regular intervals over the course of one year and Cqs were plotted versus incubation time (A&B). Regression analysis for MTB positive (C) and MTB negative (D) cartridges. The percent change between the mean Cq values at the test time point and mean Cq values at baseline [(T_test_-T_baseline_)/T_baseline_]*100 ] were plotted versus the time cartridges were incubated. The stability target is <10% different from the baseline. The 30-day timepoint was excluded due to experimental error because the amount of MTB DNA included in the test was 10X more than in the other timepoints.

Figure S1.

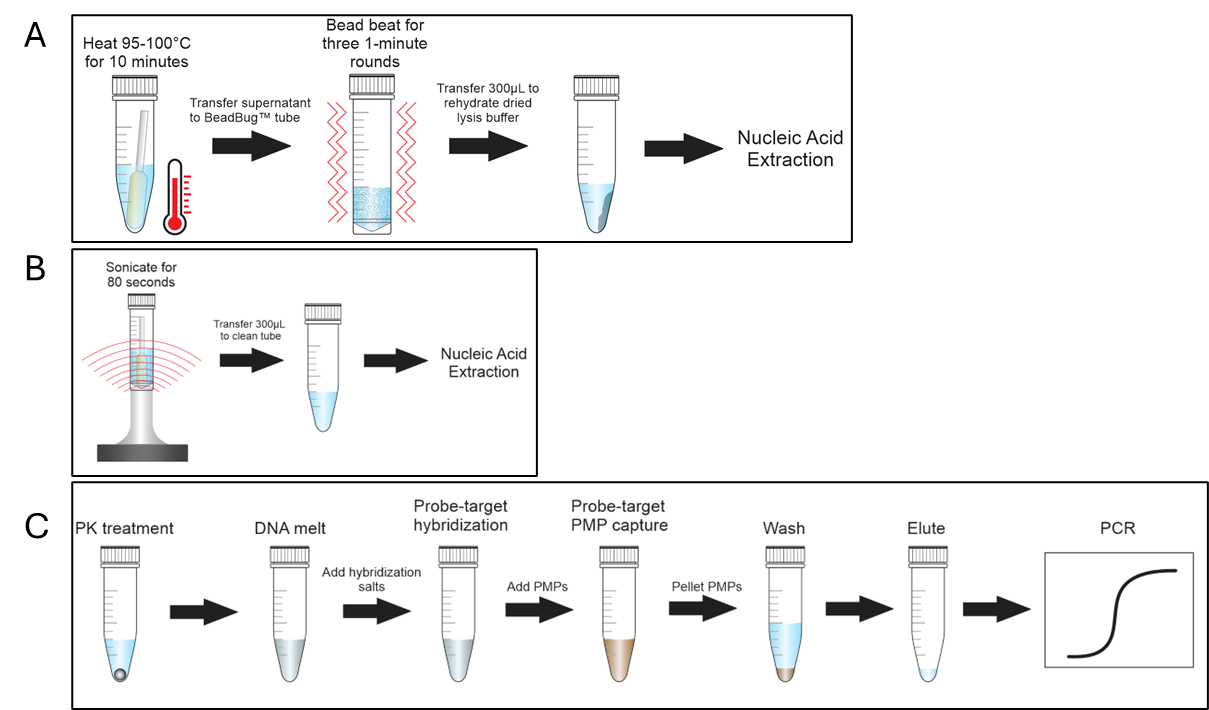

Figure S2.

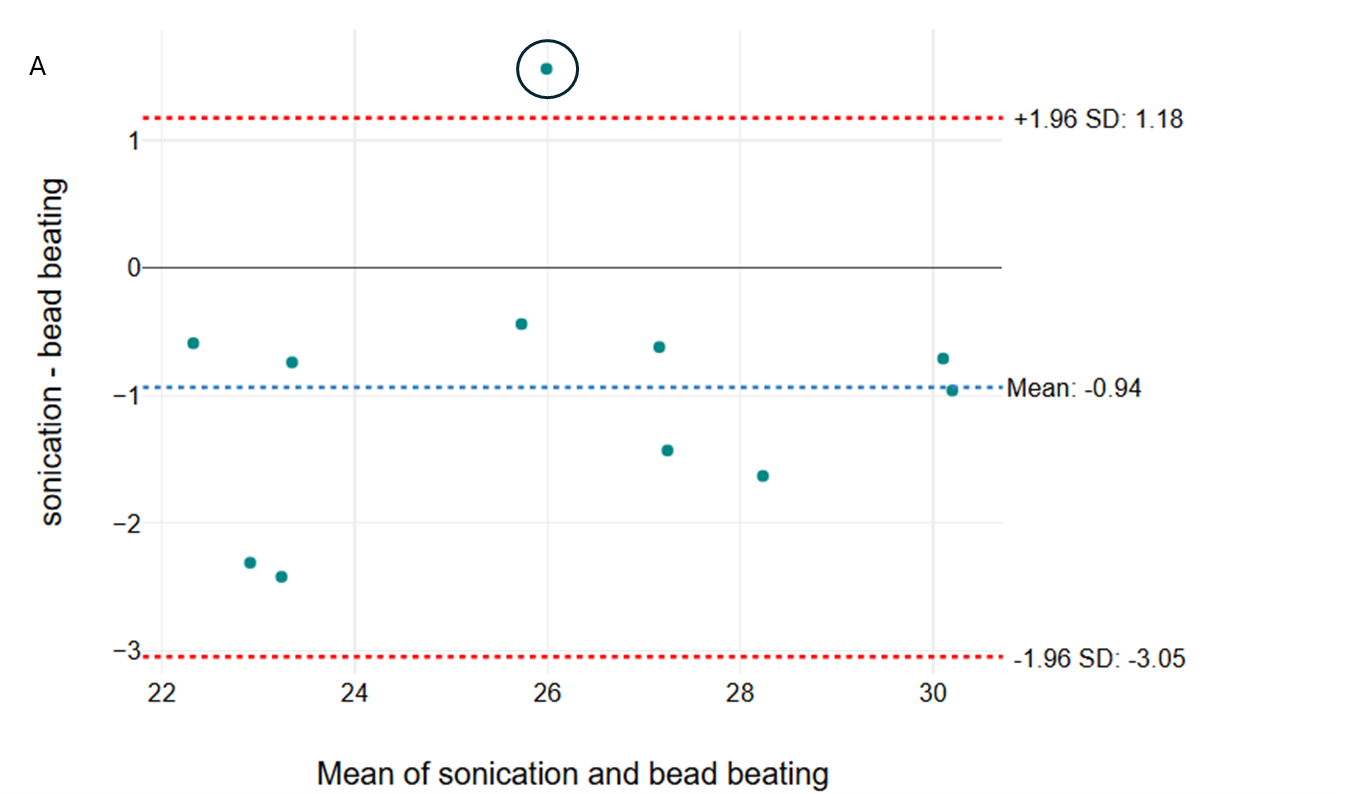

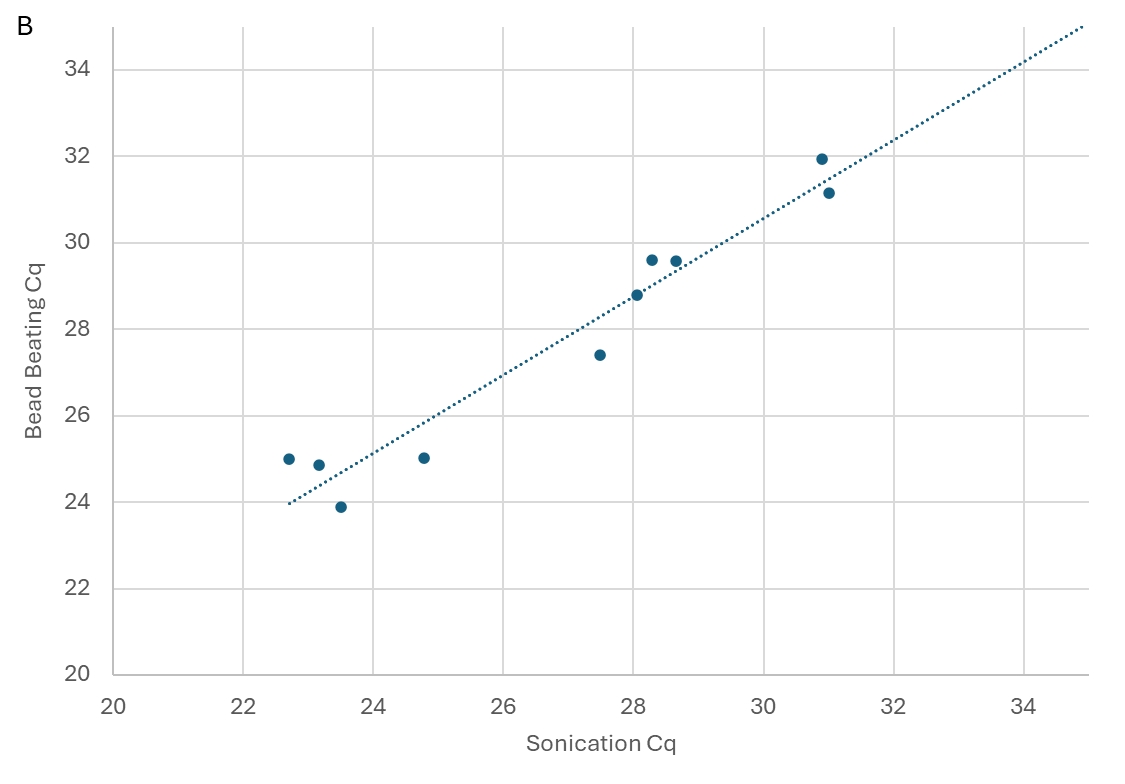

Figure S2

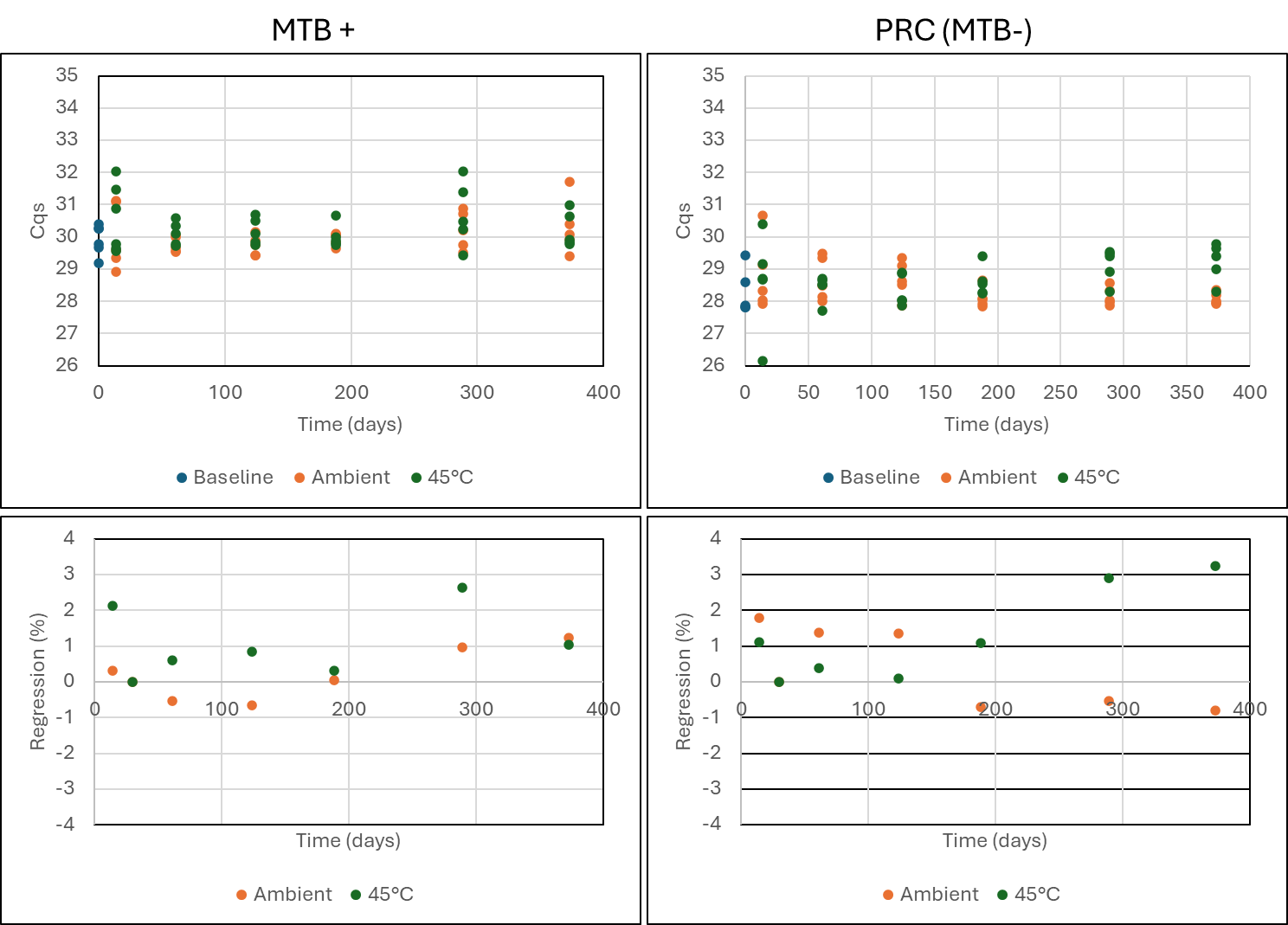

A

B

D

C
